# Prevalence of microvascular complications in Diabetes Mellitus patients attending a Nephrology outpatient department at a tertiary care hospital in Bangladesh

**DOI:** 10.1101/2025.08.12.25333485

**Authors:** Ferdous Jahan, Mohammed Anwer, Mohammad Rezaul Alam

**Affiliations:** Department of Nephrology, Bangladesh Medical University; Department of Physical Sciences, Independent University, Bangladesh

**Keywords:** Type 2 diabetes mellitus, Microvascular complications, Diabetic nephropathy, Diabetic retinopathy, Diabetic neuropathy, chronic kidney disease

## Abstract

**Background:** Type 2 diabetes mellitus (T2DM) is a chronic metabolic disorder with a rising global prevalence and represents a significant public health challenge. Persistent hyperglycemia in T2DM is closely linked to the development of microvascular complications.

**Objective:** To assess the prevalence of microvascular complications — namely diabetic nephropathy, retinopathy, and neuropathy—among patients with type 2 diabetes mellitus attending the nephrology outpatient department of a tertiary care hospital.

**Method:** This hospital-based cross-sectional study was carried out at Bangladesh Medical University, Dhaka, and included 283 confirmed cases of type 2 diabetes mellitus in patients aged 20 years and above. Participants were selected consecutively from the outpatient departments. Microvascular complications were identified using clinical assessments and diagnostic investigations.

**Results:** Of the 283 patients with type 2 diabetes mellitus, 66.4% (*n* = 188) had at least one microvascular complication. Diabetic neuropathy was the most prevalent, affecting 49.8% of patients, followed by diabetic retinopathy in 38.2% — with 19.1% of these cases showing proliferative changes. Diabetic foot was identified in 24.7% of patients. A significantly higher prevalence of microvascular complications was seen among patients with a longer duration of diabetes (*p* = 0.002), coexisting hypertension (*p* = 0.005), and more advanced stages of chronic kidney disease (*p* = 0.003). Biochemical analyses showed that patients with microvascular complications had higher mean serum creatinine levels (3.68 ± 2.35 *mg*/*dL* vs. 2.57 ± 1.41 *mg*/*dL*; *p* < 0.001) and lower eGFR values (23.40 ± 14.11 vs. 31.22 ± 14.35 *mL*/*min*/1.73 *m*^2^; *p* < 0.001) compared to those without complications. No statistically significant associations were found with age, gender, BMI, family history of diabetes, lipid profile, fasting blood glucose, or HbA1c levels.

**Conclusion:** Microvascular complications are common among type 2 diabetes patients in the nephrology outpatient setting, with neuropathy being most frequent. Their strong links to longer disease duration, hypertension, and worsening kidney function highlight the need for early screening and integrated management to slow progression and improve outcomes.

## Introduction

Diabetes mellitus (DM) is one of the most prevalent non-communicable diseases globally, with an estimated 537 million adults affected in 2021 — a number projected to rise to 783 million by 2045 (1,2). Type 2 diabetes mellitus (T2DM), a chronic metabolic condition marked by insulin resistance and sustained hyperglycemia, progressively damages multiple organ systems, particularly small blood vessels (3). T2DM represents a growing global health challenge, largely due to its strong association with microvascular complications such as diabetic nephropathy, retinopathy, and neuropathy (3). These complications are primarily driven by prolonged metabolic and hemodynamic disturbances that induce oxidative stress, inflammation, and endothelial dysfunction (4).

Diabetes-related complications are generally classified as microvascular or macrovascular (5). Among these, microvascular complications — retinopathy, nephropathy, and neuropathy — are especially common and debilitating outcomes of chronic hyperglycemia, leading to capillary and small vessel damage (4). In the United States, a comprehensive study conducted from 1988 to 2014 identified diabetic nephropathy as the leading cause of end-stage renal disease (ESRD), significantly burdening nephrology services (6,7). Diabetic retinopathy continues to be a major cause of preventable blindness worldwide (8,9). Diabetic neuropathy affects up to 50% of individuals with longstanding diabetes and may present as peripheral, autonomic, or focal nerve dysfunction, severely impacting quality of life (10). It is also a major factor in the development of diabetic foot ulcers (11,12) and lower limb amputations (13,14).

The burden of T2DM complications places substantial strain on global healthcare systems. However, the prevalence and presentation of these complications vary across regions, influenced by disparities in healthcare access, socioeconomic conditions, and control of risk factors. A multinational study involving approximately 16,000 patients from 38 countries across six regions examined the patterns of vascular complications associated with T2DM (15). A summary of the findings is presented in Table 1.

**Table 1.** Summary of some results from (33)

| Table 1. Summary of some results from (33) |  |  |  |  |  |  |
| --- | --- | --- | --- | --- | --- | --- |
| Region | Africa | Americas | SE Asia | Europe | Medeter | West Pacific |
| Sample size | 812 | 2002 | 3360 | 3479 | 2182 | 4157 |
| Duration of diabetes (years) | 6.9 | 6.2 | 4.6 | 6.6 | 5.8 | 5.1 |
| CKD (%) | 2.5 | 4.5 | 0.7 | 7.4 | 1.4 | 9.0 |
| Albuminuria (%) | 2.6 | 2.0 | 5.6 | 5.6 | 4.2 | 3.1 |
| Retinopathy (%) | 3.9 | 2.7 | 3.4 | 0.9 | 3.2 | 5.7 |
| Autonomic Neuropathy (%) | 0.1 | 0.4 | 1.2 | 0.8 | 1.1 | 1.3 |
| Peripheral Neuropathy (%) | 5.8 | 5.0 | 9.6 | 9.4 | 8.3 | 5.9 |
| Diabetic foot (%) | 0.5 | 0.5 | 1.0 | 0.4 | 0.3 | 0.2 |

It is quite evident that there is a wide variation in the regional dependency of microvascular complications T2DM. In further detailed studies, in China, national surveys identified retinopathy in 27% and nephropathy in 23%, with disparities observed between urban and rural populations (16–18).

In the Middle East, data from Saudi Arabia indicated neuropathy in 39.5%, nephropathy in 25.3%, and retinopathy in 26.6% of patients with T2DM, with poor glycemic control and obesity being prominent contributing factors (19–21). The vulnerability of Africa to T2DM can be understood by the number of investigations done in this region (19–27) In sub-Saharan Africa, an Ethiopian hospital-based study reported that 37.3% of T2DM patients had at least one microvascular complication, with retinopathy being the most prevalent (28).

In Europe, the UK Prospective Diabetes Study (UKPDS) found that nearly 39% of patients had retinopathy at the time of T2DM diagnosis, and intensive glycemic control significantly reduced the risk of developing microvascular complications (29, 30). In Germany, data from the German Diabetes Study revealed that 28% of patients already had at least one microvascular complication at diagnosis, underlining the importance of early detection (31–33).

North American data, particularly from the United States, indicate a declining trend in microvascular complications over the past two decades, though nephropathy still affects 25– 35% of patients, with notable ethnic disparities (34–36). 2018).

It has been observed, across these regions, neuropathy tends to be the most prevalent microvascular complication, followed by nephropathy and retinopathy. Common risk factors include prolonged duration of diabetes, hypertension, dyslipidemia, obesity, and inadequate glycemic control. These findings emphasize the global need for integrated diabetes care, including regular screening, patient education, and multidisciplinary management to reduce the burden of microvascular complications and improve patient outcomes.

Several factors are associated with the development of microvascular complications. These include poor glycemic control, long duration of diabetes, older age, presence of hypertension, and hyperlipidemia (24, 37). Even at the time of initial diagnosis, many patients already present with one or more complications, highlighting the silent and progressive nature of T2DM (37). Studies emphasize that early diagnosis, regular monitoring, and strict metabolic control are key strategies to prevent or delay the onset of complications (21, 24).

South Asia, including Bangladesh, is experiencing a disproportionately high increase in diabetes prevalence due to rapid urbanization, sedentary lifestyles, and dietary changes (38, 39). In Bangladesh alone, the prevalence of T2DM among adults is estimated at 9–10%, with significant public health implications (40–43). These results indicate that during a period of 2018 to 2022, the prevalence of diabetes has increased from 14% to 16.1%

Despite growing awareness, there is limited data on the specific burden of microvascular complications among diabetic patients attending nephrology outpatient departments. These patients are often at increased risk of renal involvement and may also present with other vascular complications. Understanding the prevalence and pattern of microvascular complications in this subgroup is essential for targeted screening and intervention.

In Bangladesh, the burden of diabetic nephropathy is increasingly recognized in tertiary care settings. However, there is a paucity of local data on the concurrent prevalence of other microvascular complications among diabetic patients who present to nephrology outpatient departments (OPDs). Understanding the pattern of these complications in this high-risk group is vital, as coexisting retinopathy and neuropathy often accelerate morbidity and further compromise renal outcomes (44–47).

Early detection and integrated management of microvascular complications are key to reducing the burden of diabetes-related morbidity and mortality (48). However, in Bangladesh, underdiagnosis and inadequate screening practices persist, particularly in patients who present with advanced renal disease. Identifying the magnitude and predictors of microvascular complications among patients attending nephrology OPDs can guide the development of comprehensive screening protocols and multidisciplinary management strategies.

Understanding the burden and mechanisms of these complications is essential for developing early detection strategies and integrated care models to prevent irreversible damage and improve long-term outcomes in T2DM patients. Therefore, this study aims to determine the prevalence and patterns of microvascular complications among patients with type 2 diabetes mellitus attending the Nephrology OPD in a tertiary care hospital in Bangladesh. The findings will contribute to local evidence needed to strengthen clinical practice and inform targeted interventions to improve patient outcomes.

## Material and Methods

This is a post-facto (retrospective, *a-posteriori*) study conducted at the Department of Nephrology of Bangladesh Medical University, Dhaka, between 2021 and 2023. Participants were selected from amongst the patients who came for treatment at the outpatient department. The sample size *n* was calculated using

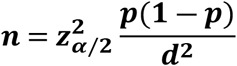

where **z**_**α**/**2**_ = **1 96** is the two-tailed 95% confidence level

*p* ≈ 0.16, the microvascular prevalence from previous studies (38, 39)

*d* = 0.05 is the error level

With these values, the sample size *n* is evaluated to be 283.

Therefore, the study included 283 adults (181 men and 102 women) aged 20 years and above with a confirmed diagnosis of type 2 diabetes mellitus. Patients with acute kidney injury, non-diabetic kidney diseases, or incomplete records were excluded. Data were collected through face-to-face interviews using a pre-structured questionnaire, along with a review of medical records and recent laboratory tests. Variables gathered included demographic information, age at diabetes onset, duration of the disease, comorbidities such as hypertension, and key biochemical markers (HbA1c, lipid profile, serum creatinine, and 24-hour urinary protein). Chronic kidney disease (CKD) stages were determined using estimated glomerular filtration rate (eGFR) derived from serum creatinine levels. Microvascular complications — including retinopathy, neuropathy, and diabetic foot were identified through clinical examination and specialist confirmation, with ophthalmologic and neurologic assessments conducted when needed. Data entry and analysis were performed using SPSS version 27. Categorical variables were analyzed with chi-square tests, while continuous variables were compared using independent t-tests. A *p*-value of less than 0.05 was considered statistically significant.

## Results and Discussion

This study provides valuable insights into the baseline characteristics, biochemical profile, and burden of microvascular complications among patients with type 2 diabetes mellitus (T2DM) attending the outpatient nephrology department in a tertiary care hospital in Bangladesh. The baseline characteristics of the patients in the sample are shown in Table 2.

**Table-2.** Baseline characteristics of patients (*n* = 283)

| Table-2: Baseline characteristics of patients ($n = 283$) | | |
| --- | --- | --- |
| Variables | Frequency | Percentage (%) |
| Age group (years) |  |  |
| < 40 | 16 | 5.7 |
| 41– 60 | 179 | 63.3 |
| > 60 | 88 | 31.1 |
| Mean ± SD | 55.7 ± 10.03 | |
| Gender |  |  |
| Male | 181 | 64.0 |
| Female | 102 | 36.0 |
| Male: Female ratio | 1.8: 1 |  |
| Socioeconomic status |  |  |
| Lower middle class | 77 | 27.2 |
| Middle class | 143 | 50.5 |
| Upper middle class | 63 | 22.3 |
| Duration of diabetes (years) |  |  |
| < 10 | 163 | 57.6 |
| 10 – 20 | 105 | 37.1 |
| > 20 | 15 | 5.3 |
| Mean ± SD | 10.5 ± 6.08 | |
| Family history of DM | 127 | 44.9 |
| Hypertension | 200 | 70.7 |
| BMI ($kg/m^2$) | | |
| Underweight | 12 | 4.2 |
| Normal weight | 186 | 65.7 |
| Overweight | 79 | 27.9 |
| Obese | 6 | 2.1 |

As shown in Table 2, the mean age of the patients was 55.7 ± 10.03 years, with the majority (63.3%) in the 41–60 years age group, and a male predominance (64%, male-to-female ratio 1.8:1). This demographic pattern is consistent with other studies conducted in Bangladesh and South Asia, which indicate that T2DM commonly affects middle-aged adults with a slight male predominance (38–41).

Notably, over half of the patients (57.6%) had a duration of diabetes of less than 10 years (Table 2), yet the prevalence of hypertension was found to be as high at 60%, indicating an early clustering of cardiovascular risk factors. This finding is comparable with other local and regional studies showing high rates of hypertension among diabetic populations (49).

In this cohort, the mean BMI was 23.49 ± 3.06 *kg*/*m*^2^ (Table 2), with 65.7% of patients having normal BMI, further supporting the well-established evidence that South Asians are more prone to T2DM and its complications at lower BMI thresholds due to increased visceral adiposity and insulin resistance (50).

The biochemical profile (Table 3) revealed a mean HbA1c of 7.25 ± 1.72%, indicating suboptimal glycemic control, slightly exceeding the target HbA1c of < 7% recommended by the American Diabetes Association (http://diabetes.org). Additionally, the mean serum creatinine was elevated at 3.31 ± 2.14 mg/dL, with a mean eGFR of ± 14.64 *mL*/*min*/1.73 *m*^2^, reflecting a substantial burden of renal impairment among these patients, which is expected in a nephrology clinic population.

**Table-3.** Biochemical and clinical profile of the patients.

| Table-3: Biochemical and clinical profile of the patients |  |  |
| --- | --- | --- |
| Variables | Mean ± SD | Range (min – max) |
| UTP24h (gm) | $4.90 \pm 21.05$ | 0.1-290.0 |
| Serum TC (mg/dl) | $169.81 \pm 55.18$ | 1.3-320.0 |
| Serum LDL (mg/dl) | $116.16 \pm 47.31$ | 28.0-288.0 |
| Serum HDL (mg/dl) | $44.20 \pm 30.37$ | 13.0-335.0 |
| Serum TG (mg/dl) | $187.96 \pm 99.98$ | 10.3-777.0 |
| FBS (mmol/L) | $8.24 \pm 3.62$ | 3.3-26.7 |
| HbA1c (%) | $7.25 \pm 1.72$ | 4.0-17.0 |
| Serum creatinine (mg/dL) | $3.31 \pm 2.14$ | 1.2-13.0 |
| BMI (kg/m<sup>2</sup>) | $23.49 \pm 3.06$ | 15.2-36.9 |
| eGFR (mL/min/1.73 m<sup>2</sup>) | $26.03 \pm 14.64$ | 3.3-69.1 |

Table 4 highlights that 66.4% of the patients had at least one microvascular complication. Diabetic neuropathy was the most prevalent (49.8%), followed by diabetic retinopathy (38.2%), including 19.1% with the proliferative form, while diabetic foot was present in 24.7% of patients. These findings are comparable to previous studies in Bangladesh and other low- and middle-income countries where neuropathy remains the most common microvascular complication in long-standing diabetes (51).

**Table-4.** Prevalence of microvascular complications among patients.

| Table-4: Prevalence of microvascular complications among patients |  |  |
| --- | --- | --- |
| Variables | Frequency | Percentage |
| Diabetic retinopathy | 108 | 38.2 |
| Proliferative Diabetic retinopathy | 54 | 19.1 |
| Diabetic neuropathy | 141 | 49.8 |
| Diabetic foot | 70 | 24.7 |
| Overall microvascular complications | 188 | 66.4 |

As shown in Table 5, microvascular complications were significantly associated with longer duration of diabetes (*p* = 0.002), presence of hypertension (*p* = 0.005), advanced CKD stages (*p* = 0.003), higher serum creatinine (*p* < 0.001), and lower eGFR (*p* < 0.001). These findings highlight the cumulative impact of poor glycemic and blood pressure control on the development of diabetic nephropathy and other microvascular complications (Adler et al., 2003).

**Table-5.**
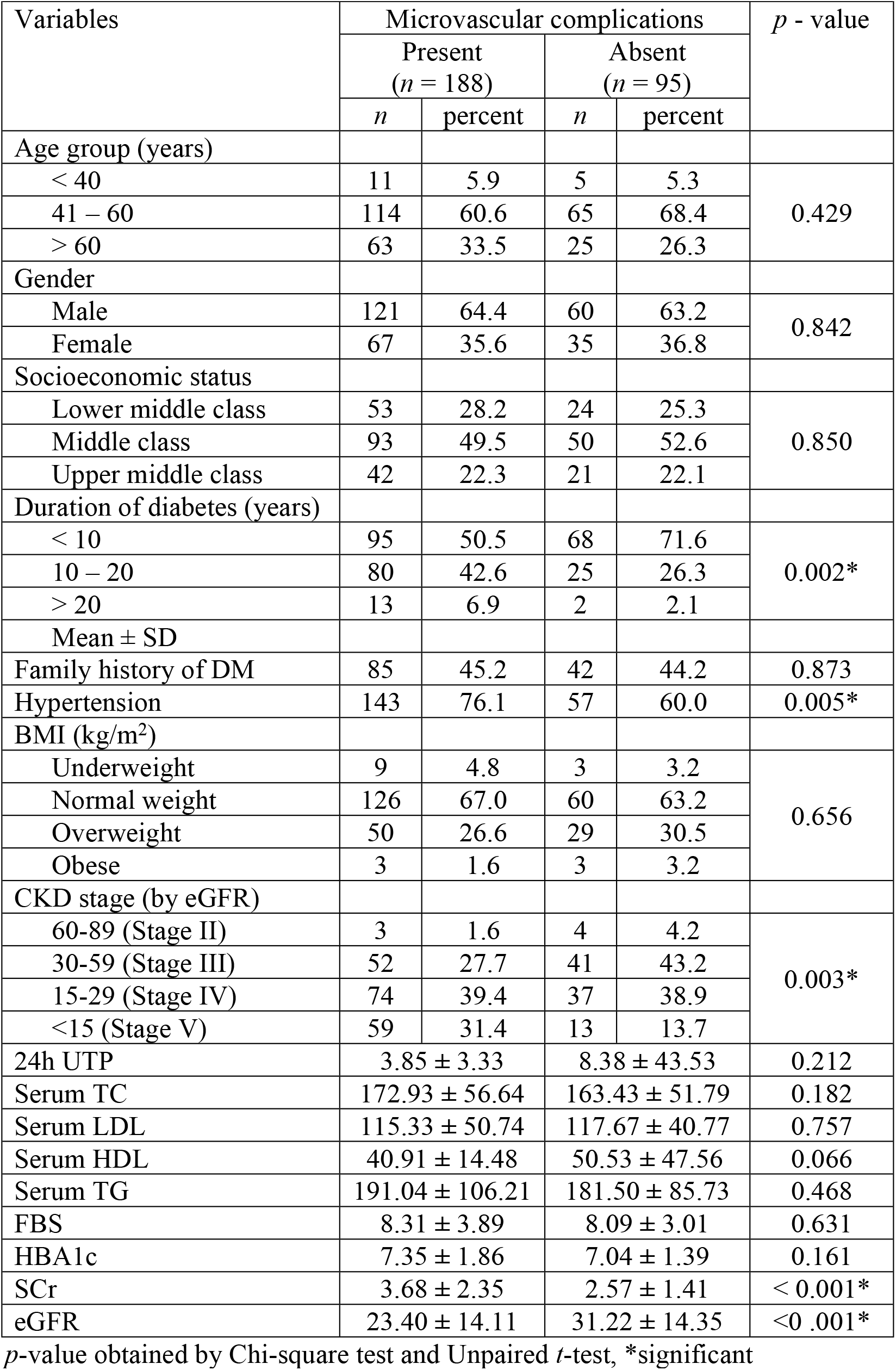
Association of microvascular complications with clinical and demographic factors among patients.

|  |  |  |  |  |  |
| --- | --- | --- | --- | --- | --- |
| Variables | Microvascular complications |  |  |  | *p* - value |
|  | Present<br>(*n* = 188) |  | Absent<br>(*n* = 95) |  |  |
|  | *n* | percent | *n* | percent |  |
| Age group (years) |  |  |  |  |  |
| < 40 | 11 | 5.9 | 5 | 5.3 | 0.429 |
| 41 – 60 | 114 | 60.6 | 65 | 68.4 |  |
| > 60 | 63 | 33.5 | 25 | 26.3 |  |
| Gender |  |  |  |  |  |
| Male | 121 | 64.4 | 60 | 63.2 | 0.842 |
| Female | 67 | 35.6 | 35 | 36.8 |  |
| Socioeconomic status |  |  |  |  |  |
| Lower middle class | 53 | 28.2 | 24 | 25.3 | 0.850 |
| Middle class | 93 | 49.5 | 50 | 52.6 |  |
| Upper middle class | 42 | 22.3 | 21 | 22.1 |  |
| Duration of diabetes (years) |  |  |  |  |  |
| < 10 | 95 | 50.5 | 68 | 71.6 | 0.002* |
| 10 – 20 | 80 | 42.6 | 25 | 26.3 |  |
| > 20 | 13 | 6.9 | 2 | 2.1 |  |
| Mean ± SD |  |  |  |  |  |
| Family history of DM | 85 | 45.2 | 42 | 44.2 | 0.873 |
| Hypertension | 143 | 76.1 | 57 | 60.0 | 0.005* |
| BMI (kg/m<sup>2</sup>) |  |  |  |  |  |
| Underweight | 9 | 4.8 | 3 | 3.2 | 0.656 |
| Normal weight | 126 | 67.0 | 60 | 63.2 |  |
| Overweight | 50 | 26.6 | 29 | 30.5 |  |
| Obese | 3 | 1.6 | 3 | 3.2 |  |
| CKD stage (by eGFR) |  |  |  |  |  |
| 60-89 (Stage II) | 3 | 1.6 | 4 | 4.2 | 0.003* |
| 30-59 (Stage III) | 52 | 27.7 | 41 | 43.2 |  |
| 15-29 (Stage IV) | 74 | 39.4 | 37 | 38.9 |  |
| <15 (Stage V) | 59 | 31.4 | 13 | 13.7 |  |
| 24h UTP | 3.85 ± 3.33 |  | 8.38 ± 43.53 |  | 0.212 |
| Serum TC | 172.93 ± 56.64 |  | 163.43 ± 51.79 |  | 0.182 |
| Serum LDL | 115.33 ± 50.74 |  | 117.67 ± 40.77 |  | 0.757 |
| Serum HDL | 40.91 ± 14.48 |  | 50.53 ± 47.56 |  | 0.066 |
| Serum TG | 191.04 ± 106.21 |  | 181.50 ± 85.73 |  | 0.468 |
| FBS | 8.31 ± 3.89 |  | 8.09 ± 3.01 |  | 0.631 |
| HBA1c | 7.35 ± 1.86 |  | 7.04 ± 1.39 |  | 0.161 |
| SCr | 3.68 ± 2.35 |  | 2.57 ± 1.41 |  | < 0.001* |
| eGFR | 23.40 ± 14.11 |  | 31.22 ± 14.35 |  | <0 .001* |

*p*-value obtained by Chi-square test and Unpaired *t*-test, \*significant

Interestingly, BMI and most lipid parameters (TC, LDL, TG) were not significantly associated with microvascular complications (Table 4). This could be due to overall suboptimal metabolic control among most patients, masking individual contributions of isolated lipid abnormalities. These findings emphasize that addressing all modifiable risk factors – including hyperglycemia, hypertension, and dyslipidemia – is critical to prevent or delay microvascular damage (http://diabetes.org).

The high prevalence of complications shown in Tables 4 and 5 underscores the need for systematic early screening, especially for neuropathy and retinopathy, and integrated multidisciplinary care targeting tight glycemic and blood pressure control, lipid management, and renal function preservation (https://kidgo.org).

## Conclusion

In summary, this study demonstrates a high prevalence of microvascular complications among T2DM patients attending a nephrology OPD in a tertiary care hospital in Bangladesh. Duration of diabetes, hypertension, and worsening renal function were significantly associated with microvascular complications. These findings underscore the need for multidisciplinary interventions focusing on early detection and comprehensive management to prevent progression and improve patient outcomes.

**Limitations** of this study include its single-center design and cross-sectional nature, which restrict causal inference. However, the findings provide baseline data to guide larger, multicenter longitudinal studies exploring the burden and determinants of microvascular complications in the Bangladeshi diabetic population.

## Data availability statement

The data is proprietary of the hospital where the study was conducted. However, the data can be made available upon request.

## Ethics statement

All patients consented to participate in the study with anonymity. The IRB of Bangabandhu Sheikh Mujib Medical University (BSMMU, predecessor of Bangladesh Medical University, BMU) waived the ethical requirement citing ‘no patients were selected for the purpose of research, but rather their past medical record were used after they received regular treatment.’

## Author contributions

FJ: Conceptualization, Data collection, Data curation, Formal analysis, Investigation, Methodology, Resources, Writing – original draft, Writing – review & editing. MRA: Conceptualization, Data collection, Data curation, Formal analysis. MA: Methodology, Software Supervision, Validation, Writing – review & editing.

